# Does explainable AI-ECG heart age differentiate pathological from physiological LV remodeling? A multi-cohort analysis including young elite athletes

**DOI:** 10.1101/2025.09.06.25335008

**Authors:** Philip Hempel, Tabea Steinbrinker, Lennart Graf, Srushhti Trivedi, Bjørn-Jostein Singstad, Mark Abela, David Niederseer, Marcus Vollmer, Marcus Dörr, Nicolai Spicher, Dagmar Krefting

**Author notes:** Shared last authorship; corresponding author.

## Abstract

**Aim:** Artificial intelligence applied to electrocardiography (AI-ECG) can derive a *heart age* or *ECG-age*, potentially reflecting waveform patterns that indicate cumulative myocardial stress. The heart age gap (HA-gap, Δ_*age*_) is defined as the difference between a person’s ECG-age and chronological age. Former studies suggest a threshold of Δ_*age*_ *>* 8 yrs as a biomarker for accelerated biological age, associated with higher risk for cardiovascular events. In this study, we investigate whether Δ_*age*_ differentiates training-induced physiological from pathological left ventricular remodeling.

**Methods:** An AI-ECG was applied to 162 resting 12-lead ECGs of each professional footballers, population controls without cardiovascular disease, and patients with systolic heart failure (HF). Explainable AI identified contributing leads and waveforms, and results were compared with established ECG voltage criteria for left ventricular hypertrophy (Sokolow–Lyon, Cornell) and low QRS voltage (LQRSV).

**Results:** Accelerated HA (Δ_*age*,+_) was present in 38.9% of athletes, 35.8% of community controls, and 96.9% of HF patients. As a diagnostic criterion, accelerated HA achieved 96.9% sensitivity and 62.7% specificity for distinguishing diseased from healthy cohorts. In contrast, classical ECG voltage criteria showed lower sensitivity (6–17%) but higher specificity (85–100%). Correlation analyses confirmed significant associations of HA-gap with Cornell voltage (*ρ* = 0.25, *p <* 0.001) and LQRSV (limb: *ρ* = *−*0.43, *p <* 0.001; precordial:*ρ* = *−*0.32, *p <* 0.001).

**Conclusions:** The AI-based HA-gap is a multi-factorial marker of ventricular remodeling beyond mass and can separate benign athletic hypertrophy from pathological remodeling with high sensitivity. Incorporating athlete and youth cohorts into model development could further improve specificity to enable future application in preventive and sports cardiology.

## Introduction

Cardiologists use established electrocardiography (ECG) risk markers such as repolarisation abnormalities (e.g. ST-segment depression, T-wave inversion), depolarisation or conduction-delays (e.g. wide or fragmented QRS), and voltage surrogates such as the Sokolow–Lyon (SL) index to identify patient at risks that need further medical attention. In recent years, artificial intelligence (AI) has redefined automated ECG analysis by showing strong predictive power for mortality and cardiac disease.[1, 2, 3, 4] Furthermore, emerging risk markers based on AI-ECG can capture overall cardiovascular health rather than a single cardiac condition. One such marker is ECG-based *heart age* or *ECG-age*, which is calculated from multifactorial waveform signatures linked to broader physiology and the concept of *biological age*[5, 6]. *The difference between chronological age and ECG-age can be denoted as heart age gap (HA-gap, Δ*_*age*_). A Δ_*age*_ *>* 8 years has been associated with increased risks of mortality, heart failure (HF), and atrial fibrillation, and was validated in Brazil, the US, and Germany[5, 7, 6], indicating that these effects generalize across diverse health care systems and populations. Since ECG-age approximates biological heart age, it appears at least partly modifiable: lower values have been linked to favorable lifestyles such as regular exercise, a cardioprotective diet, and optimal risk-factor control, whereas higher values are associated with detrimental exposures including hypertension, diabetes, smoking, or air pollution.[5, 8, 7]

Therefore, ECG-age may serve as a novel risk marker in groups where traditional screening is limited. Young individuals and athletes are often underrepresented in population cohorts, and thereby also during development of AI-ECG models, yet may benefit most from early detection of accelerated or abnormal cardiac aging.[9, 10, 11] Notably, elite athletes constitute only 0.0025% of the general population,[12] raising the question of whether current AI-ECG models can reliably distinguish benign training-induced physiological from pathological disease-related ventricular remodeling, thereby avoiding false positives while still detecting individuals at risk.

In athletes, left ventricular hypertrophy (LVH) is common and often leads to larger/broader ECG signals caused by the increased muscle mass and complicate ECG interpretation [13, 8, 14, 11]. However, LVH can not only be caused by physiological adaptation to training [13, 15], but also result from pathological pressure or volume overload. In contrast, myocardial fibrosis is expected only in pathologic hypertrophy since the replacement of active muscle tissue with electrically not active binding tissue is necessary to limit the dilation of the myocardium. This process slows the electrical conduction and may attenuate the ECG signal [16]. All these structural processes unfold over years or decades, leaving electrical signatures that AI-ECG models can detect long before clinical symptoms appear [7, 6]. While most findings in athletes are generally benign, they may overlap with features of pathological hypertrophy and fibrosis, as seen in systolic HF, where ECG changes carry significant prognostic implications.[17] Notably, peri-pubertal “juvenile” ECG variants, particularly anterior T-wave inversion, can mimic cardiomyopathic patterns in adolescent athletes, raising concerns about the performance of AI-ECG in this group. Preliminary work indicates that only age-aware AI can distinguish benign maturational changes from arrhythmogenic right ventricular cardiomyopathy (ARVC).[18, 19]

Therefore, in this study we applied the external validated AI-ECG model to predict Δ_*age*_ for elite athletes, patients with systolic HF, and matched community controls without cardiovascular diseases (CVD) to investigate its performance in these diverse cohorts in comparison with established ECG parameters (SL, Cornell, LQRSV). Furthermore, we used explainable AI (XAI) to identify the leads and waveform segments that contributed most to the ECG-age, i.e. are used by the AI-ECG model to quantify the underlying cardiac aging effects. The central goal of this analysis was to enlighten the “black box” nature of AI and provide interpretable evidence for the recognized patterns. To this purpose, we also correlate Δ_*age*_ to the established ECG parameters to indicate which are also represented by the AI-ECG.

## Methods

We conducted a cross-sectional, multicohort analysis to test whether ECG-age can discriminate physiological remodeling in athletes, that are mainly based on LVH, from pathological remodeling by HF-driven by adaptions like fibrosis. Resting 12-lead ECGs were assembled from three cohorts representing distinct cardiac phenotypes: (i) elite *Athletes* from Spain’s top football league (*La Liga* (mixed-sport, intermittent high-intensity training), (ii) age- and sex-matched *Population controls* from the community-based Study of Health in Pomerania (SHIP), and (iii) *Systolic HF controls* from MIMIC-IV that include hospitalized patients only. All athletes and population controls were asymptomatic, reported no relevant family history of inherited cardiac disease, and had no cardiovascular diagnoses, thereby serving as healthy controls (true negative). Each athlete was matched to a community control and to an HF patient by sex and age using optimized assignment by the Hungarian algorithm [20]). For both MIMIC-IV and SHIP, only one ECG per patient was included in the analysis (subjects were excluded from the pool after one ECG matched by age and sex). All cohort studies adhered to the ethical guidelines of the 1964 Helsinki declaration. All participants provided written informed broad consent before participation in the studies. Baseline characteristics of the cohorts are presented in Table 1. Technical details on ECG acquisition (including recording devices, sampling rates, formats, and processing) are provided in the Supplementary Table 1.

**Table 1.** Baseline characteristics by dataset. Cohorts: SHIP (Population control, *n* = 162), LaLiga (Athletes, *n* = 162), and MIMIC (systolic HF control, *n* = 162). Accelerated HA (Δ_*age*,+_) defined as Δ_*age*_ *>* 8 years. All participants are male; ECG voltage thresholds are male-specific.

| Dataset | Baseline | | ECG criterion, $n$ (%) | | | | |
| --- | --- | --- | --- | --- | --- | --- | --- |
| | Age<br>mean ± SD (y) | HR<br>mean ± SD (bpm) | Accelerated<br>HA ($\Delta_{age,+}$) | SL<br>>3.5 mV | Cornell<br>>2.8 mV | LQRSV<br>limb < 0.5 mV | LQRSV<br>prec < 1.0 mV |
| SHIP | 26.1 ± 4.6 | 66.3 ± 11.6 | 58 (35.8%) | 13 (8.0%) | 2 (1.2%) | 0 (0.0%) | 0 (0.0%) |
| LaLiga | 25.7 ± 5.1 | 54.3 ± 9.5 | 63 (38.9%) | 35 (21.6%) | 27 (16.7%) | 0 (0.0%) | 0 (0.0%) |
| MIMIC | 25.8 ± 5.1 | 84.7 ± 22.9 | 157 (96.9%) | 22 (13.6%) | 27 (16.7%) | 15 (9.3%) | 9 (5.6%) |

### LaLiga

The dataset contains 163 ECGs from 54 male football players between 17–37 years from a single team participating in the Spanish top-tier professional football league “La Liga”. The cohort represents a population of elite male athletes competing at the UEFA Pro level. All available 12-lead resting ECGs were collected as part of standardized annual medical examinations conducted by team medical staff between 2018 and 2022. Recordings took place at five time points: the 2018–2019 postseasons and the pre-seasons of 2019, 2020, and 2021 [21]. One recording could not be processed due to technical issues, resulting in 162 ECGs for analysis.

### SHIP

The Study of Health in Pomerania (SHIP) is a population-based cohort study in north-eastern Germany. Participants were examined at baseline (SHIP-TREND-0, T0) and at a 5-year follow-up (SHIP-TREND-1, T1) between 1997 and 2012.

For this analysis, SHIP was restricted to participants from T0 and T1 without cardiovascular disease or risk factors at either exam and with an age between 20–37 years. Exclusion criteria included: (i) a diagnosis of atrial fibrillation, HF, myocardial infarction, or hypertension; (ii) prescription of cardiovascular or antidiabetic medication (ATC codes C02, C03, C07, C08, C09, C07AA, C08CA, C01AA, A10A, A10BA); (iii) a positive family history of myocardial infarction or stroke; and (iv) cardiovascular mortality during follow-up. These criteria yielded a healthy subset of 2628 ECGs from 1828 individuals. Notably, the minimum age in SHIP was slightly higher than in LaLiga (20 vs. 17 years), which we slightly balanced by age/sex matching by highly penalizing this rare matches with factor 10 [20]. However, the mean ages and SD are similar over all subjects and datasets (see Table 1)

### MIMIC-IV

The MIMIC-IV–ECG dataset [22, 23] was acquired from adult patients admitted to Beth Israel Deaconess Medical Center between 2008 and 2019. For this study, we selected ECGs from patients between 18 and 37 years diagnosed with systolic HF, as identified in the MIMIC-IV-ECG-Ext-ICD dataset [24, 25, 26] by ICD-10 codes I50.20, I50.21, I50.22, I50.23, I50.40, I50.41, I50.42, and I50.43. 1, 640 ECGs from individuals under the age of 37 were sampled to match the elite athletes by age and sex.

#### ECG Processing

All ECG recordings were processed using a harmonized, automated pipeline. For each ECG, the raw waveforms were first loaded from their respective source formats (see Supplementary Table 1). Subsequently, signals were reordered into standard 12-lead format and, if necessary, converted to the standard mV unit, including alignment of ECGs to the isoelectric line by subtraction of the mean value of each lead’s signal. All recordings were recorded in resting, standard supine position. For each dataset, recordings were resampled to a target sampling rate of 400 Hz and symmetrically padded to a uniform length of 4096 samples (10.2 seconds).

#### AI-ECG Model

ECG-age was obtained using a previously developed and externally validated artificial intelligence model based on a one-dimensional residual neural network [5]. The model analyses standard 12-lead ECG waveforms (10-second recordings, sampled at 400 Hz; 4096 samples per lead) and directly provides an ECG-age, without requiring manual feature extraction or conventional ECG parameters. The HA-gap (Δ_*age*_) was defined as the difference between ECG-age and chronological age. A positive Δ_*age*_ indicates that the electrical pattern of the heart appears “older” than expected for the patient’s chronological age, whereas a negative Δ_*age*_ suggests a “younger” appearing ECG. In line with previous studies, we categorized heart age status into three groups: *regular HA* (Δ_*age,∼*_) when |Δ_*age*_| *≤* 8 years, *accelerated HA* (Δ_*age*,+_) when Δ_*age*_ *>* 8 years, and *decelerated HA* (Δ_*age,−*_) when Δ_*age*_ *< −*8 years. An accelerated HA has been proposed as a marker of biological aging beyond chronological age and is associated with increased cardiovascular risk [5, 7, 6].

#### XAI analysis

We used the Captum library’s implementation of the Integrated Gradients algorithm [27], a widely adopted method for highlighting which parts of the ECG signal most influenced the model’s prediction, ECG-age respectively. The algorithm compares the model output and the internal ECG processing when the input signal is present versus when it is gradually removed, thereby estimating the contribution of each data point to its outcome.

Two levels of explanation were derived: “Lead importance” was calculated as the relative contribution of each of the 12 ECG leads, normalized so that all leads together sum to 100%. For visualization, we focused on the precordial leads (V1– V6), since they capture most of the voltage and repolarization changes relevant to left ventricular remodeling [6]. “Waveform importance” was assessed within a cardiac cycle by aligning signals to the R-peak [6] and mapping contributions onto the main ECG segments (P-wave, QRS-complex, T-wave). Positive values indicate ECG waveforms that increased the predicted ECG-age, while negative values decreased it.

#### ECG parameters

We assessed classical ECG indices including SL/Cornell voltage and LQRSV (limb and precordial) as surrogates of LVH and myocardial remodeling [15, 28, 29]. R-peaks were detected using a consensus pipeline across all 12 leads to ensure robust amplitude measurements [6]. Associations between HA-gap and ECG indices were quantified using Spearman correlation, with *p <* 0.05 considered significant. For descriptive purposes, scatterplots were generated to illustrate distributions and pairwise associations stratified by cohort and HA status. Also, the resting heart rate was calculated for all datasets based on the R-peak information. In addition, we tested accelerated HA (Δ_*age*,+_ *>* 8 years) against the classic ECG criteria for discriminating healthy (SHIP and LaLiga) from diseased (MIMIC systolic HF) cohorts. For each ECG criterion (SL, Cornell, LQRSV limb/precordial, and accelerated HA), we calculated sensitivity, specificity, positive predictive value (PPV), and negative predictive value (NPV) with 95% Wilson confidence intervals. Baseline prevalence is summarized in Table 1.

All code used for this publication and outlined in the methods is made openly available to enable further research, reproducibility, and clinical interpretability: https://gitlab.gwdg.de/medinfpub/biosignal-processing-group/xai_in_sport_medicine

## Results

### Overall performance

Across cohorts, the mean HA-gap was modest in healthy groups (SHIP: +6.6 years; LaLiga: +6.9 years) and markedly higher in systolic HF (MIMIC: +35.3 years). Accelerated HA (Δ_*age*,+_) occurred in 35.8% of SHIP participants, 38.9% of LaLiga athletes, and 96.9% of systolic HF patients (Table 1, Fig. 2). Decelerated HA (Δ_*age,−*_) was rare (*n* = 1 in LaLiga) and excluded from downstream voltage analyses.

**Fig. 1.**
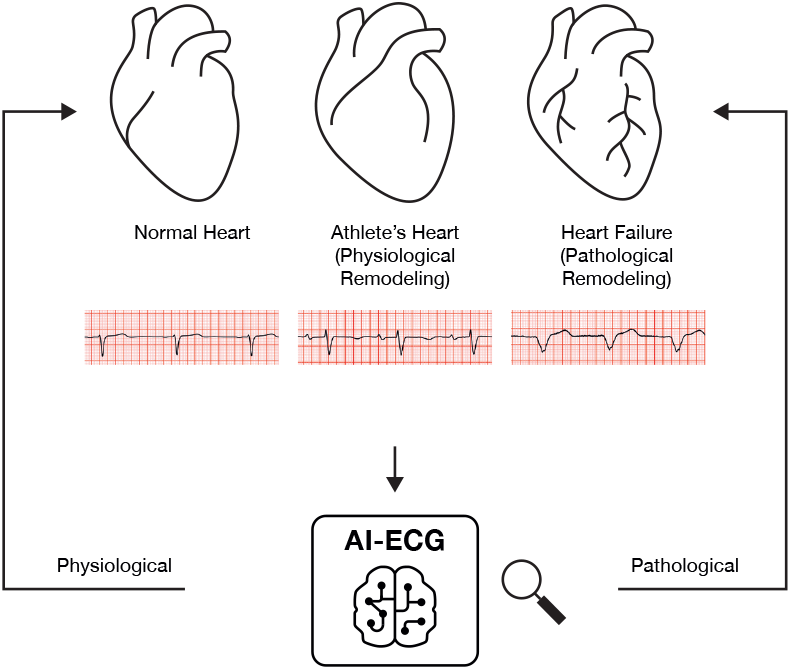
Graphical abstract. Athlete-centered comparison against two comparators: negative controls (age/sex-matched healthy community adults) and a disease-positive control (systolic HF). ECG-age shows modest HA-gap in athletes/healthy youth but strong HA-gap in HF; explainability differs by cohort (V4 vs V1), and traditional LVH criteria do not discriminate physiologic from pathologic remodeling; abbreviations: heart age gap (HA-gap).

**Fig. 2.**
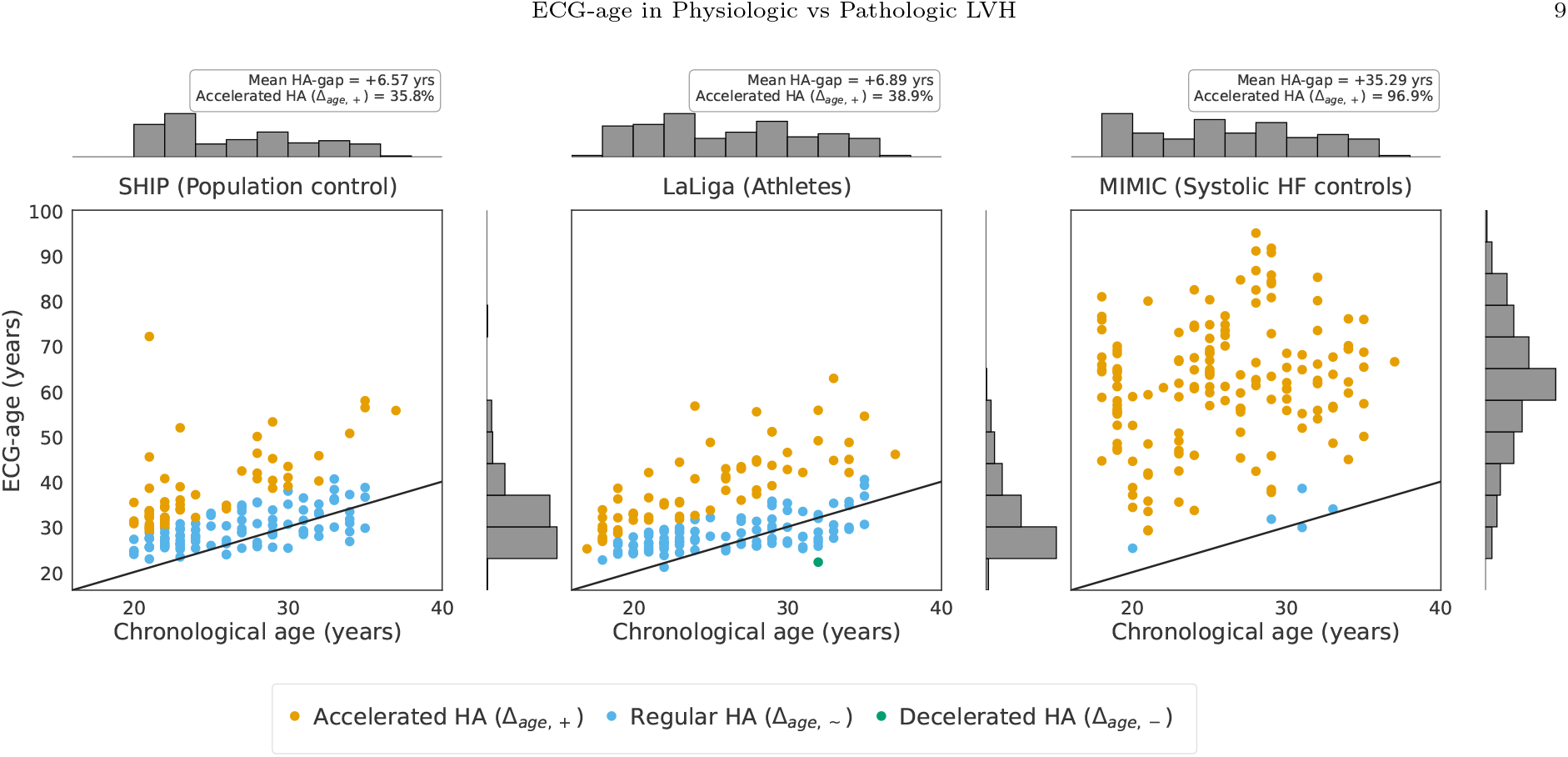
ECG-age versus chronological age in three matched cohorts of *n* = 162. Each point represents one 10-s, 12-lead ECG; colours indicate HA status (accelerated, regular, decelerated). The 1 : 1 reference line visualizes the HA-gap as the vertical distance (Δ_*age*_ = ECG-age *−* chronological age). Mean HA-gap was modest in SHIP (+6.6 years) and LaLiga athletes (+6.9 years), but markedly higher in systolic HF (MIMIC-IV, +35.3 years). Accelerated HA occurred in 35.8%, 38.9%, and 96.9% of SHIP, LaLiga, and HF, respectively. Normality of paired differences was rejected for SHIP vs LaLiga (Shapiro–Wilk *W* = 0.982, *p* = 0.031), so a Wilcoxon signed-rank test was applied (*p* = 0.836). For SHIP vs HF (*W* = 0.988, *p* = 0.166) and LaLiga vs HF (*W* = 0.986, *p* = 0.112), assumptions were met and paired *t*-tests showed significantly larger HA-gaps in HF versus both groups (both *p <* 0.001).

### XAI

Lead importance (Fig. 3) concentrated in the mid-precordial lead V4 in SHIP (Population control) and LaLiga (Athletes), while in MIMIC (Systolic HF control) it shifted toward V1.

**Fig. 3.**
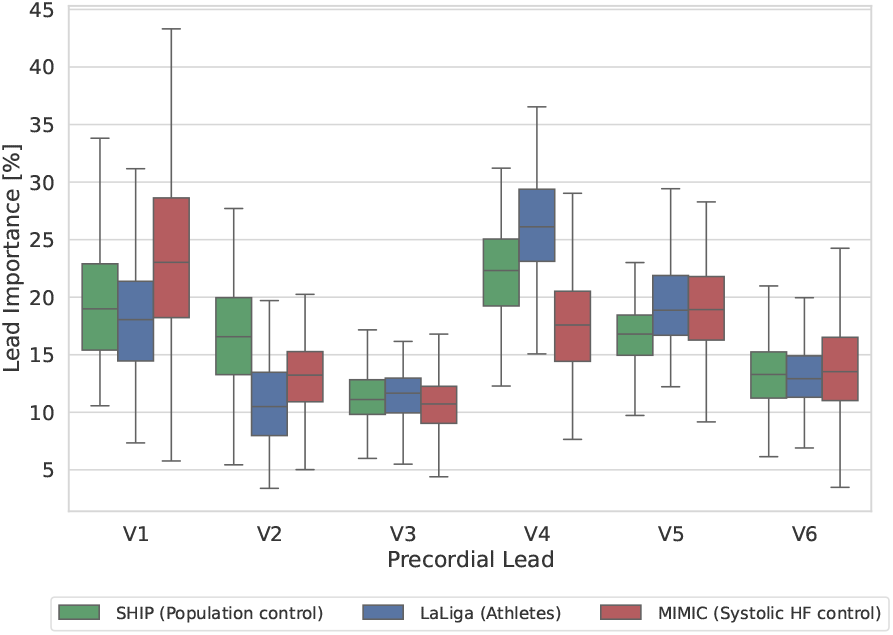
Lead-wise importance across the precordial leads (V1–V6). Values indicate each lead’s share of the model’s contribution to ECG-age (sum across all 12 leads equals 100%). SHIP (Population control) and LaLiga (Athletes) concentrate in V4, whereas MIMIC (Systolic HF control) shifts toward V1 with a broader distribution. Limb leads are omitted for clarity.

Waveform importance (Fig. 4) showed that in V4 of both SHIP and LaLiga the QRS-complex and T-wave were important for the AI-ECG, with LaLiga displaying a stronger and slightly shifted T-wave contribution. In MIMIC, T-wave importance in V4 was minimal. In V1, the XAI results showed no P-wave contribution for LaLiga, moderate in SHIP, and biphasic in MIMIC.

**Fig. 4.**
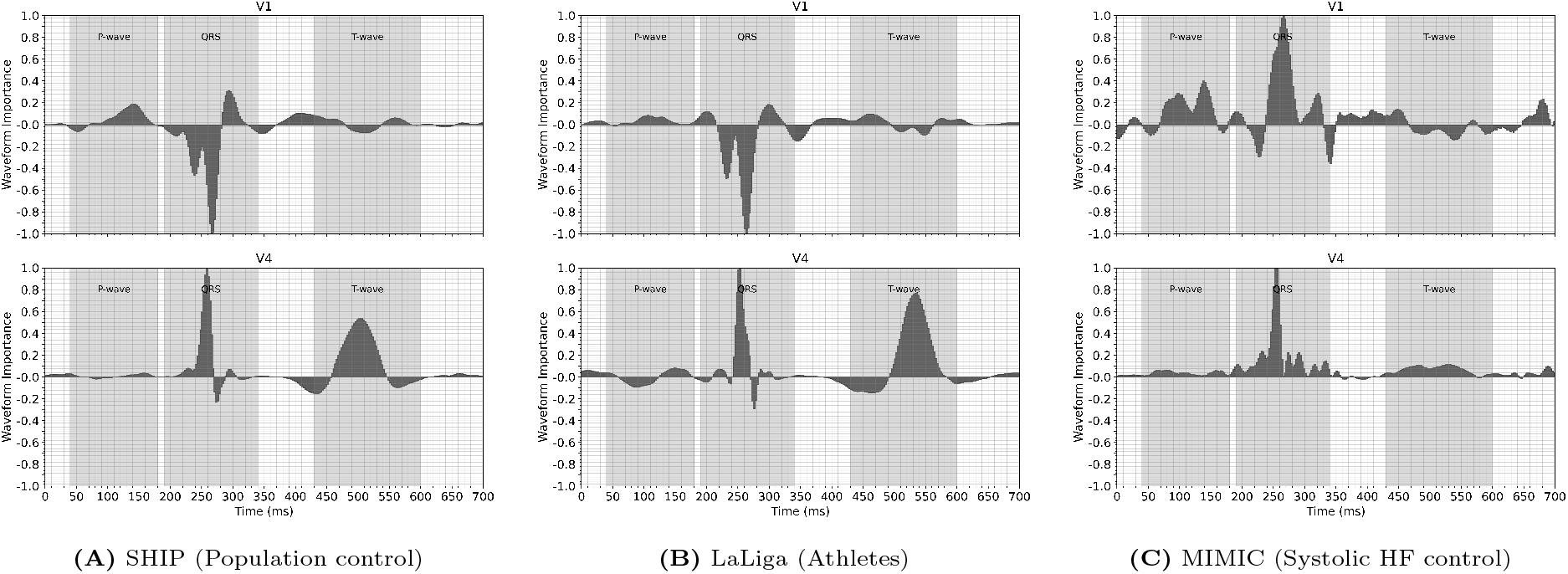
The waveform importance indicates how a particular part of the ECG waveform contributed to the ECG-age. It was calculated by normalized integrated-gradients XAI relevances for the precordial leads and overlaid via R-peak over all ECGs and heartbeats; shaded windows indicate P-wave, QRS-complex, and T-wave. Positive values indicate higher ECG-age while negative values point to ECG parts resulting in lower ECG-age. Panels show cohort means: (A) SHIP (Population control), (B) LaLiga (Athletes), (C) MIMIC (Systolic HF control). In SHIP, a moderate P-wave relevance is seen in V1; it is minimal in LaLiga but highest in MIMIC, suggesting greater atrial contribution. For ventricular depolarization, SHIP shows predominantly negative QRS relevance in V1 (features pushing predictions younger), whereas MIMIC exhibits broad positive QRS relevance in V1 with an RSR-like pattern, consistent with septal conduction delay/diffuse remodeling. In V4, SHIP and LaLiga show positive QRS/T relevance—compatible with physiological voltage/repolarization—while MIMIC shifts attention away from mid-precordial leads. T-wave (repolarization) relevance is higher in LaLiga than in SHIP and low in MIMIC, underscoring differences between physiological and pathological remodeling.

### Classic ECG parameters

Baseline characteristics was summarized in Table 1. Mean age was comparable across cohorts, whereas resting heart rate differed as expected, being lowest in LaLiga (Athletes) and highest in MIMIC (Systolic HF control). Classic voltage parameters were less frequent in SHIP (Population controls; SL 8.0%, Cornell 1.2%, LQRSV 0%) compared to LaLiga (Athletes; SL 21.6%, Cornell 16.7%, LQRSV 0%) and MIMIC (Systolic HF controls; SL 13.6%, Cornell 16.7%, LQRSV 9.3% limb and 5.6% precordial). Diagnostic performance is shown in Table 2. Classic voltage parameters had high specificity (85–100%) but low sensitivity (6–17%). In contrast, accelerated HA achieved 96.9% sensitivity, 62.7% specificity, with a PPV of 56.5% and an NPV of 97.6% for distinguishing HF from healthy cohorts. Scatterplots in Fig. 5 illustrate the association between HA-gap and voltage parameters across cohorts. Significant correlations were observed with Cornell voltage (*ρ* = 0.25, *p <* 0.001), SL voltage (*ρ* = *−*0.22, *p <* 0.001), LQRSV (limb, *ρ* = *−*0.43, *p <* 0.001), and LQRSV (precordial, *ρ* = *−*0.32, *p <* 0.001).

**Table 2.** Diagnostic performance of classic ECG voltage criteria and accelerated HA (defined as Δ_*age*,+_ *>* 8 years) as ECG criterion for distinguishing healthy (SHIP, *n* = 162; LaLiga, *n* = 162) from diseased (MIMIC, systolic HF control, *n* = 162) cohorts. Values are given as percentage with 95% Wilson confidence intervals.

| ECG criterion | Sensitivity | Specificity | PPV | NPV |
| --- | --- | --- | --- | --- |
| SL LVH | 13.6% [9.1, 19.7] | 85.2% [80.9, 88.6] | 31.4% [21.8, 43.0] | 66.3% [61.7, 70.7] |
| Cornell LVH | 16.7% [11.7, 23.2] | 91.0% [87.4, 93.7] | 48.2% [35.7, 61.0] | 68.6% [64.1, 72.8] |
| LQRSV (limb) | 9.3% [5.7, 14.7] | 100.0% [98.8, 100.0] | 100.0% [79.6, 100.0] | 68.8% [64.5, 72.8] |
| LQRSV (precordial) | 5.6% [2.9, 10.2] | 100.0% [98.8, 100.0] | 100.0% [70.1, 100.0] | 67.9% [63.6, 72.0] |
| Accelerated HA ($\Delta_{age,+}$) | 96.9% [93.0, 98.7] | 62.7% [57.3, 67.7] | 56.5% [50.6, 62.2] | 97.6% [94.5, 99.0] |

**Fig. 5.**
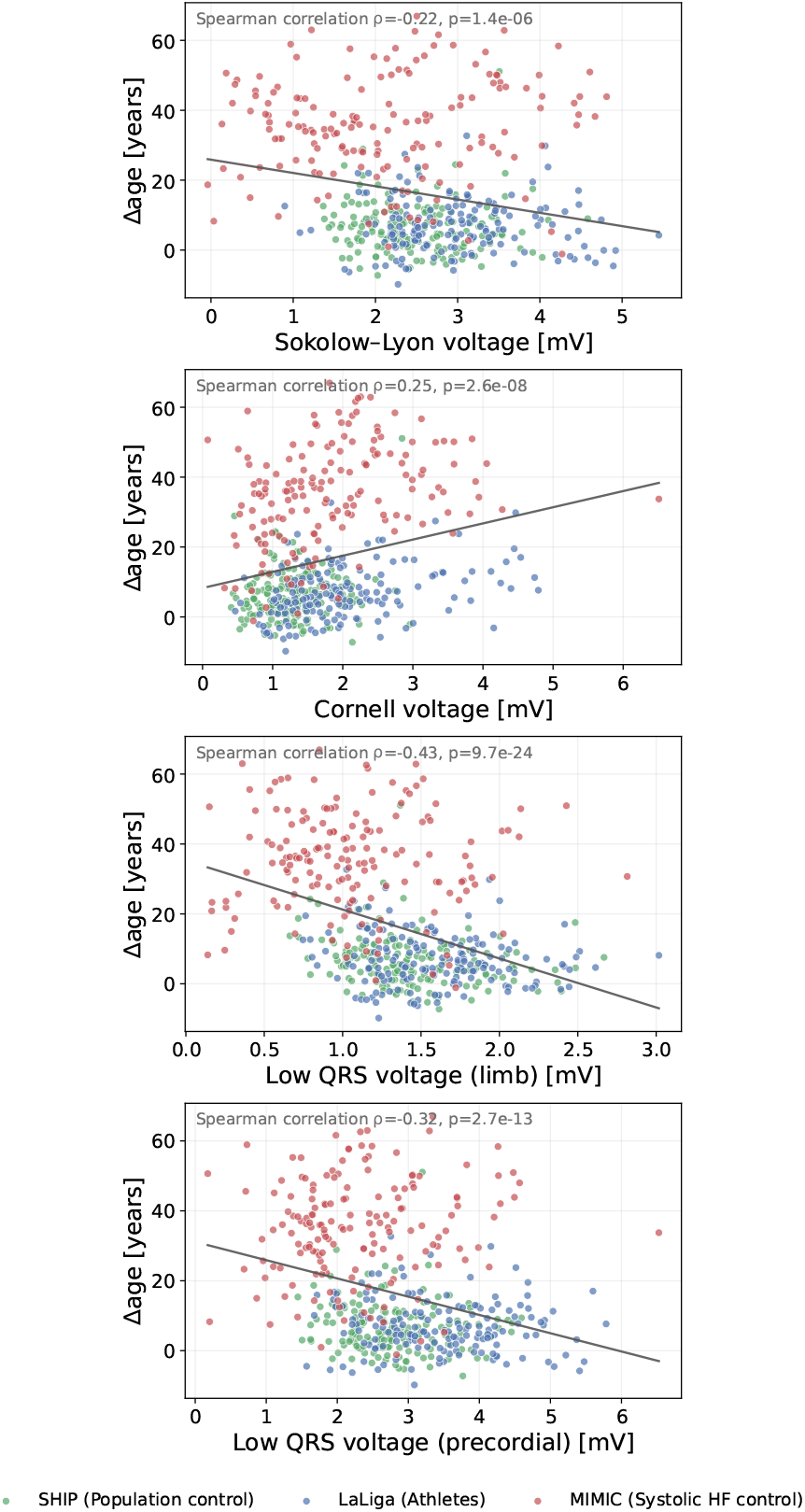
Scatterplots of classic ECG voltage parameters against HA-gap, (Δ_*age*_) across SHIP (Population control), LaLiga (Athletes), and MIMIC (Systolic HF control). Panels display (top to bottom): SL voltage, Cornell voltage, LQRSV (limb), and LQRSV (precordial). Shapiro–Wilk tests indicated non-normal error distributions in SHIP (*W* = 0.896, *p <* 0.001) and LaLiga (*W* = 0.977, *p* = 0.007), but not in MIMIC (*W* = 0.990, *p* = 0.351); therefore, associations were assessed using Spearman rank correlation. Significant correlations were observed: Cornell voltage (*ρ* = 0.25, *p <* 0.001), LQRSV (limb, *ρ* = *−*0.43, *p <* 0.001), LQRSV (precordial, *ρ* = *−*0.32, *p <* 0.001), and SL voltage (*ρ* = *−*0.22, *p <* 0.001).

## Discussion

In this multi-cohort study, we evaluated an AI-based ECG-age across three distinct populations: a general population (SHIP, Germany), elite footballers (LaLiga, Spain), and patients with systolic HF (MIMIC-IV, USA). Our findings highlight both the promise and limitations of ECG-age as a non-invasive biomarker for cardiac remodeling across physiological and pathological spectrums.

### AI-ECG performance and biological relevance

First, ECG-age in athletes is similar to population controls, indicating that physiological hypertrophy due to intensive training does not, in itself, lead to accelerated HA. This indicates that benign remodeling in athletes is not systematically misclassified by the AI-ECG model.

Second, in patients with systolic HF, nearly all individuals demonstrated a markedly increased HA-gap, with a mean deviation of *∼*35 years. This consistent finding underscores that accelerated HA reflects pathological remodeling, likely driven by diffuse fibrosis, chamber dilation, and impaired conduction [6].

Third, an additional observation was that approximately one-third of population controls also exhibited accelerated HA. This proportion is unexpectedly high and indicate a concrete risk that physiological remodeling in these healthy cohorts is labeled as “accelerated HA”. Used as a screening tool without clinical context, such potential misclassifications could trigger unnecessary follow-up or affect eligibility decisions in sports cardiology [14].

A central cause could be a mismatch between the patient data used for model-development and intended screening populations. Hospital-based training cohorts are typically older, have more diseases, and under-represent athletes and very young individuals. The AI-ECG model, for instance, was developed on ECG data from clinical patients with a mean age of 51[5, 6]. Thresholds and calibration derived from such data may not transfer to youth, athletes, or under-represented sex/ethnic strata, leading to systematic bias [30, 31].

### XAI: lead- and waveform-importance patterns

XAI highlighted coherent, cohort-specific lead- and waveform-importance patterns (Figs. 3–4 At the lead-level, SHIP (Population control) and LaLiga (Athletes) concentrated importance in V4, whereas MIMIC (Systolic HF control) shifted toward V1 which may indicate to left-lateralized hypertrophy in physiological remodeling versus septal/depolarization changes in pathological remodeling [13, 17]. At the waveform-level, SHIP/LaLiga showed prominent QRS and T-wave contributions in V4, with athletes exhibiting a slightly stronger and mildly shifted T-wave contribution which both could indicate that LVH is partially recognized by the AI-ECG. In MIMIC, broad positive QRS contributions were observed in V1 with minimal T-wave contribution in V4, consistent with slower ventricular conduction and attenuated repolarization[6]. P-wave contributions were minimal in LaLiga, moderate in SHIP (V1), and highest in MIMIC, indicating that atrial electrical remodeling predominantly augments ECG-age in HF. Overall, these lead-/waveform-level findings are consistent with previous finding [6] and potentially indicate that the AI-ECG model differentiate known physiological from pathological differences, including the potential role of juvenile/anterior T-wave inversion [18].

### ECG parameters

Classical ECG parameters related to the ECG-age but were not redundant with it (Fig. 5; Tables 1, 2). Cornell voltage correlated positively with Δ_*age*_ (*ρ* = 0.25, *p* < 0.001), while SL correlated negatively (*ρ* = *−*0.22, *p* < 0.001), indicating that LVH partially contribute to accelerated HA. Interestingly, LQRSV also correlated negatively (limb *ρ* = *−*0.43, precordial *ρ* = *−*0.32, both *p <* 0.001), consistent with reduced myocardial mass and/or fibrosis attenuating QRS amplitudes and manifesting as an “older” ECG-age [28, 29]. This highlights that nearly opposite voltage behaviors can belong to a structural continuum: LVH surrogates like Cornell emphasize increased depolarization amplitude typical for hypertrophy, including benign, training-related remodeling in athletes, whereas LQRSV emphasizes weakened signals compatible with reduced mass and/or fibrotic change [13, 28, 29, 16]. This opposition helps explain why HA-gap, learned from raw waveforms, can be more sensitive: the AI-ECG integrates multiple, partially competing substrates (hypertrophy-related high voltage and fibrosis-related low voltage) into a single multifactorial signature rather than relying on a single amplitude threshold [6]. In athletes, SL was frequent yet not associated with increased HA-gap supporting its potentially benign nature [13], whereas higher Cornell voltage appeared more often in athletes with accelerated HA, suggesting partial overlap between Cornell-related features and those the model interprets as age-advancing which align with contemporary LVH guidance [32]. Using the established *±*8 year threshold for HA classification [5, 7, 6], accelerated HA (Δ_*age*,+_ *>* 8 years) distinguished HF from healthy youth/athletes with **96.9% sensitivity** and **62.7% specificity** (PPV 56.5%, NPV 97.6%) (Table 2); by contrast, SL, Cornell, and LQRSV remained highly specific but comparatively insensitive [28, 29]. In practice, a profile based on lead-/waveform-importance (V1-shifted, QRS-dominated vs. V4-centric QRS/T) in combination with classic ECG parameters could possibly offer a pragmatic check when developing and reviewing AI-ECG models for screening purposes in younger/athletic cohorts [6, 14].

### Limitations

This study has several limitations. Voltage-based ECG criteria (SL and Cornell) were used as surrogates of left-ventricular mass, although their correlation with imaging-defined hypertrophy is modest and their sensitivity limited [32, 13]. We lacked longitudinal follow-up and echocardiographic data, precluding exclusion of subclinical disease and outcome validation. The sample comprised only males and the athletes were originated from a single professional football team, which restricts generalizability and prevents analysis of sex-specific differences in remodeling and ECG-age. Furthermore, the systolic HF controls were hospital-acquired, whereas population controls and athletes were outpatient screening ECGs that represent different acquisition context and comorbidity burden. In addition, the HA-gap threshold of *±*8 years was derived in older clinical cohorts and may be suboptimal for younger and athletic populations, indicating a need for recalibration. Overall, without imaging we cannot definitively distinguish physiological from pathological remodeling at the individual level. These considerations underscore the need for future sex-balanced, prospective, multi-modal studies and age-/sport-specific calibration to improve specificity in athletic and young subjects.

## Conclusion

In this multi-cohort analysis of young males, ECG-age differentiated pathological from physiological remodeling at the cohort level: controls with systolic HF showed clearly higher HA-gaps than athletes and population controls, while specificity in the healthy cohorts remained only moderate. Cohort-specific lead and waveform patterns were physiologically plausible and point to indicators for athlete- and age-specific calibration. For sports cardiology, these findings support ECG-age as an interpretable AI-based ECG criterion to identify individuals who may merit imaging or closer follow-up. Moving forward, prospective studies that integrate ECG with echocardiography, refine thresholds for youth and athletic populations, and report transparent subgroup performance are likely to determine whether AI-ECG can improve screening and longitudinal monitoring in sport medicine.

## Supporting information

supplement_deidentified

## Acknowledgments

The authors gratefully acknowledge the computing time granted by the Resource Allocation Board and provided on the supercomputer Emmy/Grete at NHR-Nord@Göttingen as part of the NHR infrastructure. The calculations for this research were conducted with computing resources under the project nhr nib00042.

## Data availability

This study analyzed ECGs from three independent cohorts:

- LaLiga (elite athletes): Publicly available ECG dataset PF12RED, collected from a Spanish professional football team and described by Muñoz et al. [21]. Available at: https://github.com/dradolfomunoz/PF12RED.
- SHIP (Study of Health in Pomerania): Population-based ECGs from north-eastern Germany. Due to participant consent, these data cannot be made publicly available. Access can be requested through a data application form at: https://fvcm.med.uni-greifswald.de.
- MIMIC-IV-ECG: De-identified hospital ECGs from the Beth Israel Deaconess Medical Center, distributed via PhysioNet as part of the MIMIC-IV project [22, 23]. Available at: https://physionet.org/content/mimic-iv-ecg/.

## Code availability

All scripts for preprocessing, matching, model inference (ECG-age), XAI (Integrated Gradients), and analysis are openly available at: https://gitlab.gwdg.de/medinfpub/biosignal-processing-group/xai_in_sport_medicine

## Author contributions statement

**P.H**.: Conceptualization, Methodology, Software, Validation, Formal Investigation, Data Curation, Visualization, Writing - Original Draft. **T.S**.: Methodology, Visualization, Writing - Review and Editing. **L.G**.: Methodology, Resources, Software, Validation, Data Curation, Writing - Review and Editing. **S.T**.: Methodology, Ethical Analysis, Writing - Review and Editing. **B.J.S**.: Resources, Writing - Review and Editing. **M.A**.: Formal Investigation, Writing - Review and Editing. **D.N**.: Formal Investigation, Writing - Review and Editing. **M.D**.: Resources, Writing - Review and Editing. **M.V**.: Resources, Formal Investigation, Data Curation, Writing - Review and Editing. **N.S**.: Formal Investigation, Methodology, Supervision, Project Administration, Resources, Writing Original Draft, Writing-Review and Editing. **D.K**.: Conceptualization, Methodology, Formal Investigation, Supervision, Funding Acquisition, Project Administration, Writing - Review and Editing.

All authors have read and approved the final manuscript.

