## supplement_deidentified for "Does explainable AI-ECG heart age differentiate pathological from physiological LV remodeling? A multi-cohort analysis including young elite athletes"

### Supplementary Information for: Does explainable AI–ECG heart age differentiate pathological from physiological LV remodeling?

Philip Hempel, Tabea Steinbrinker, Lennart Graf, Srushhti Trivedi, Bjørn-Jostein Singstad, Mark Abela, David Niederseer, Marcus Vollmer, Marcus Dörr, Nicolai Spicher*, Dagmar Krefting*

#### 1 Supplementary Tables

**Supplementary Table 1** ECG recording devices, file formats, and processing tools for each dataset.

| **Dataset** | **Recording device** | **File format** | **Tool** |
|---|---|---|---|
| LaLiga | GE USB-CAM 14 | XML | xml.etree |
| SHIP | Welch–Allyn CardioPerfect | EDF | MNE |
| MIMIC–IV | Burdick/Spacelabs, Philips, GE | WFDB | WFDB |

#### 2 Supplementary Figures

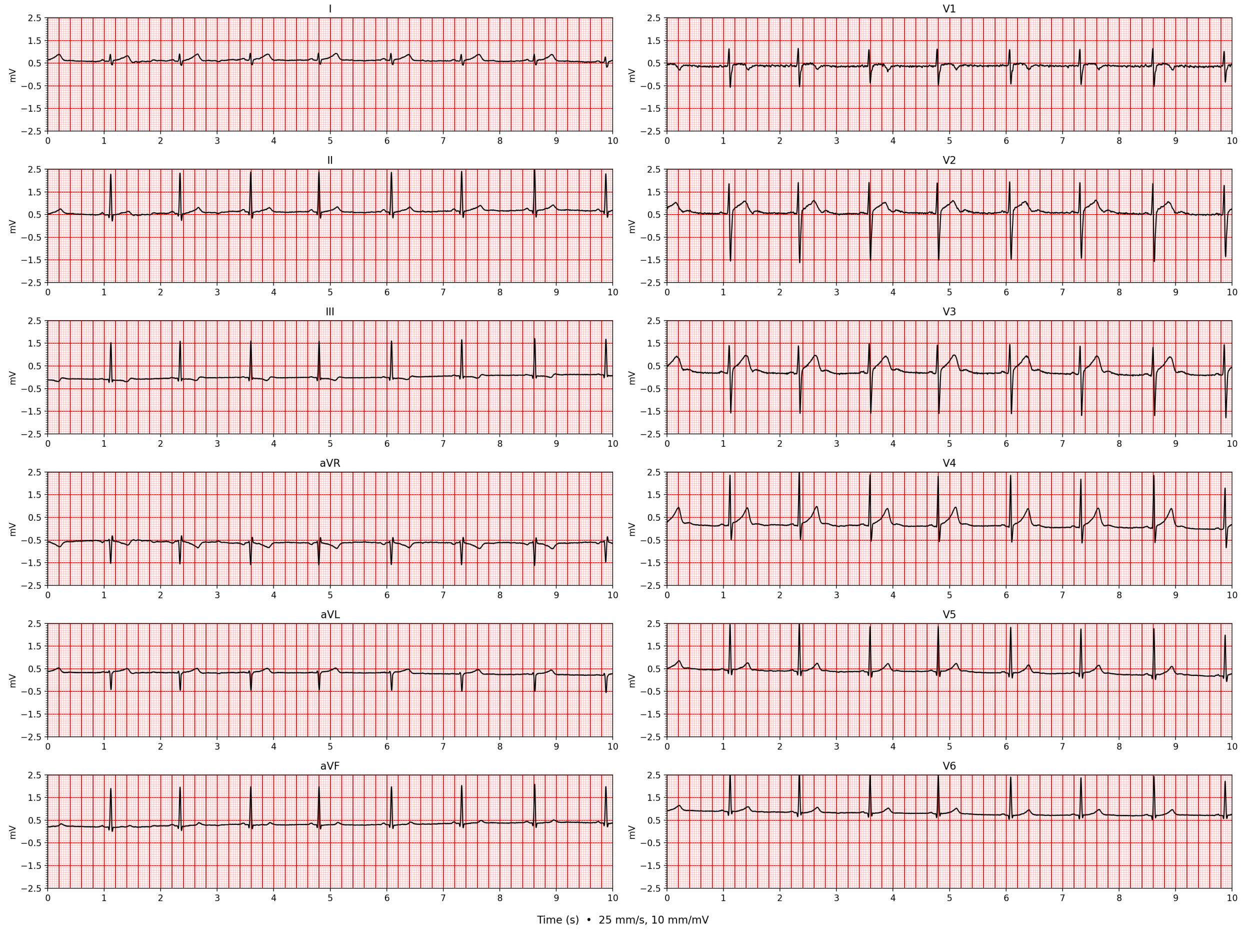


**Supplementary Figure 1:** Example of a 12 lead ECG derived from a LaLiga subject (Athlete).

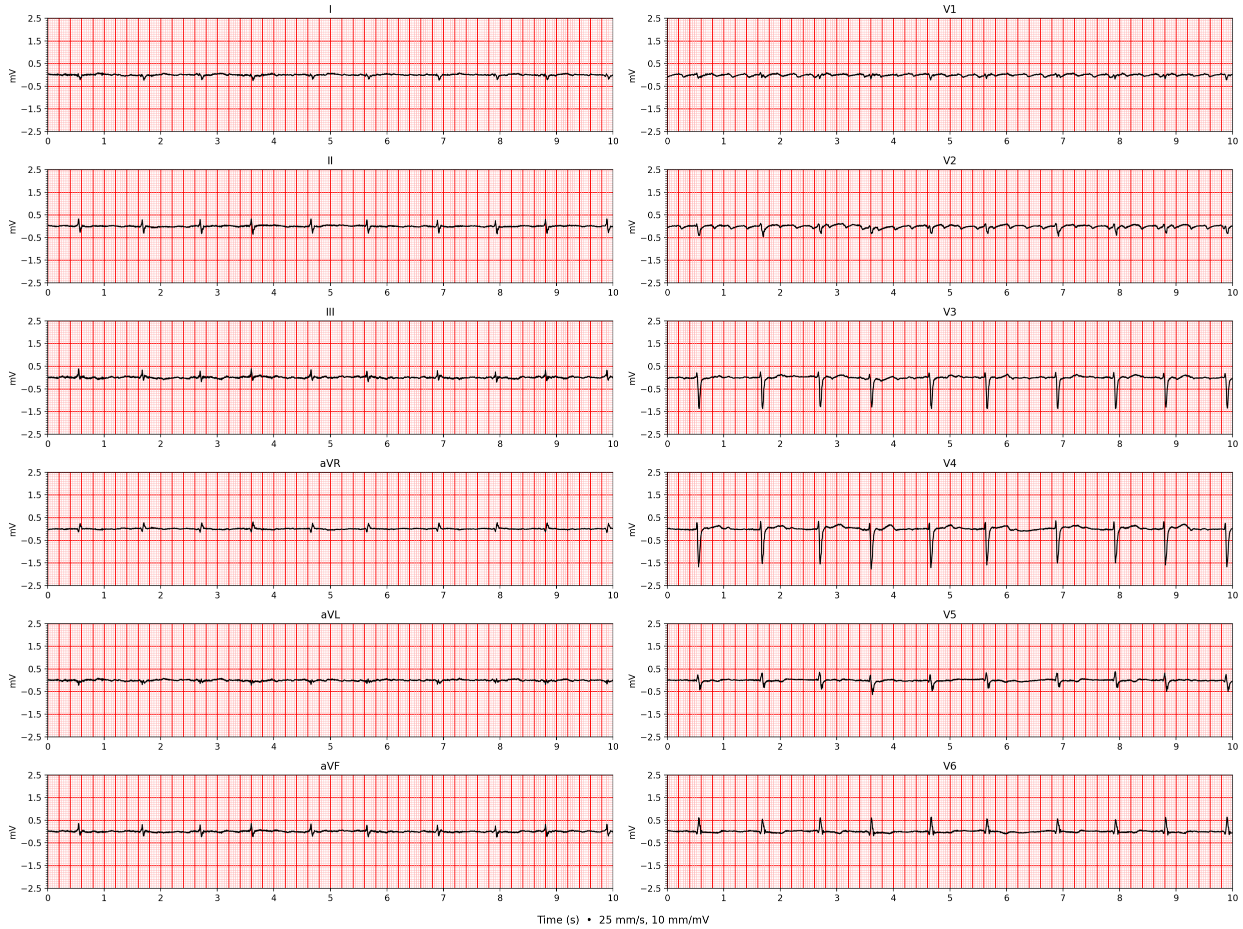


**Supplementary Figure 2:** Example of a 12 lead ECG derived from a MIMIC patient (Systolic HF control).

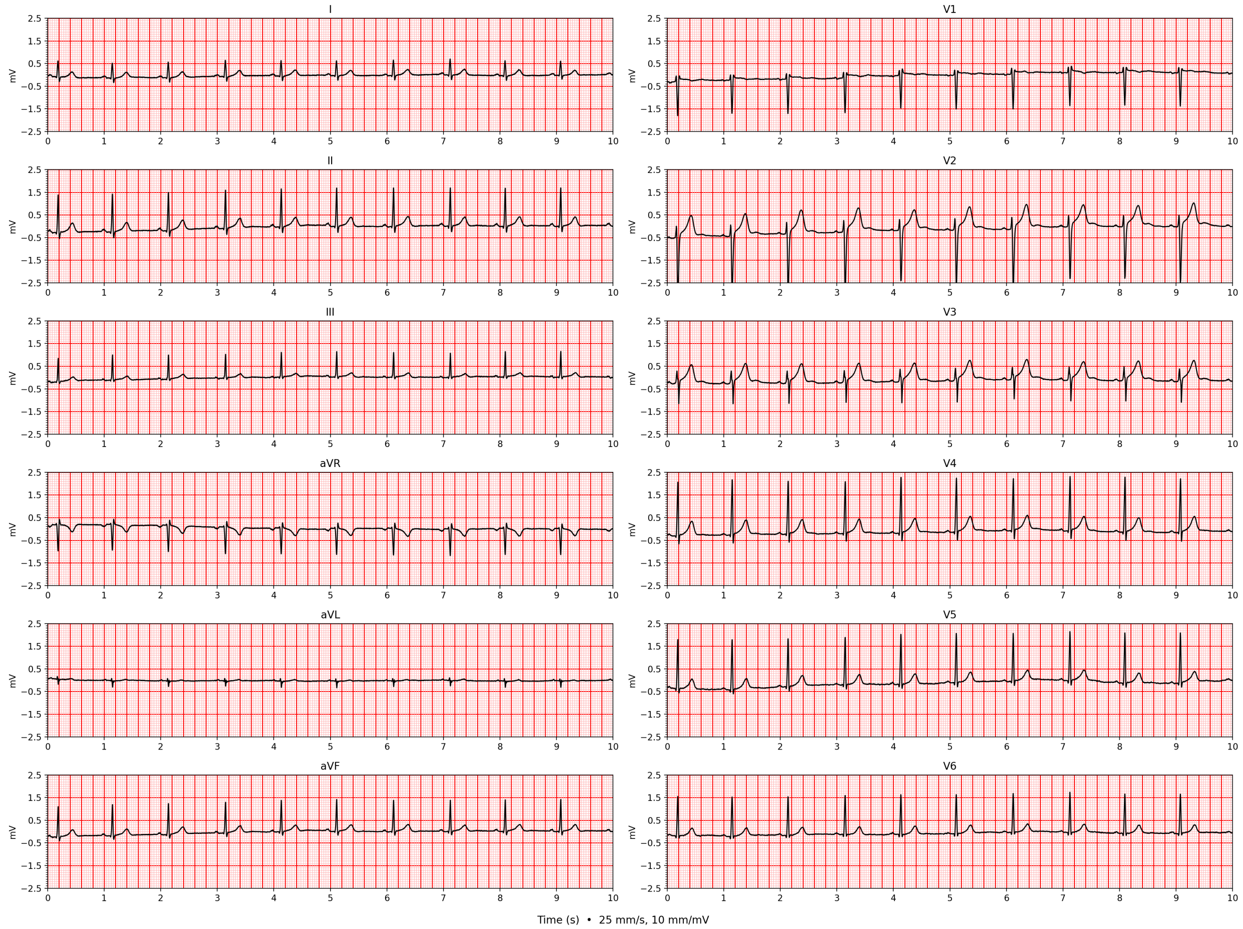


**Supplementary Figure 3:** Example of a 12 lead ECG recorded in a SHIP participant (Population control).
